# Development and Validation of a Modular Footwear Setup for testing the isolated biomechanical effects of footwear features

**DOI:** 10.64898/2026.03.30.26349729

**Authors:** Hadi Sarlak, Kamran Shakir, Giulia Rogati, Giorgia Sartorato, Alberto Leardini, Lisa Berti, Paolo Caravaggi

**Affiliations:** Department of Biomedical and Neuromotor Sciences, Alma Mater Studiorum - Università di Bologna, 40136 Bologna, Italy; (K.S.); (H.S.); (L.B.); Movement Analysis Laboratory and Functional Evaluation of Prostheses, IRCCS Istituto Ortopedico Rizzoli, Via di Barbiano 1/10, 40136 Bologna, Italy; (G.R.); (A.L.) (P.C.); Independent Researcher; (G.S.); Physical Medicine and Rehabilitation Unit, IRCCS Istituto Ortopedico Rizzoli, 40136 Bologna, Italy

**Keywords:** modular footwear, in-shoe pressure, lower-limb kinematics, therapeutic shoes, midsole, accuracy, repeatability

## Abstract

The effects of specific footwear features on biomechanical parameters are often confounded by simultaneous changes in other shoe conditions, making it difficult to identify the isolated effect of material and design properties on relevant biomechanical outcomes. This study aimed to propose a tool, namely the Modular Footwear Setup (MFS), to assess the effects of midsole modifications on lower limb joint kinematics and in-shoe plantar pressure. The MFS uses a micro-hook-and-loop fastening system and a custom alignment device to enable fast, strong, and reliable midsole attachment/detachment to/from the upper.

Accuracy and repeatability of the MFS in replicating the biomechanical outcomes of a control shoe featuring the same upper and midsole were tested in 10 healthy participants (5M,5F; age=33.2±9.2 yrs; BMI=21.5±2.8 kg*m^-2^). Participants were asked to walk wearing both the MFS and the standard control shoe in three sessions. Kinematics of lower limb joints were measured via inertial measurement units, while capacitive pressure insoles were used to measure in-shoe plantar pressure. Intraclass correlation coefficient (ICC) was used to assess the repeatability of kinematic and pressure measurements between sessions.

Statistical Parametric Mapping analysis did not identify significant differences in joint kinematics between conditions. While the MFS exhibited slightly lower peak pressure at the rearfoot, pressure parameters were not statistically different in the other foot regions. The MFS demonstrated good-to-excellent inter-session repeatability (ICC 0.84–0.97) for peak and mean pressure. Participants reported similar levels of comfort and stability in both shoes.

The findings of the present study suggest the MFS has the potential to be a reliable and accurate tool for evaluating the effect of midsole features on relevant biomechanical parameters. This modular approach may improve data-driven footwear design by providing a consistent platform for testing the effects of midsole designs and materials across various applications, including therapeutic, safety, and athletic shoes.

## Introduction

Footwear protects the foot from injury and is made using a variety of materials and shapes to address different applications and environments. There are significant differences in requirements and thus in the footwear design and materials according to the specific application. For instance, safety shoes are large, heavy footwear that can withstand high loads, protect the feet against impacts, and help isolate the body from electrical shocks (1). Sport shoes, on the other hand, are generally light and designed to improve performance. In the healthcare sector, therapeutic shoes are designed to address lower limb and foot ailments. In the post-surgery period, orthopedic shoes such as forefoot offloading shoes, are designed to protect the operated region from excessive loading (2).

Footwear modifies lower-limb kinetics and kinematics with respect to barefoot conditions (3), and its mechanical properties (e.g., sole stiffness, rocker geometry) are known to affect plantar pressure distribution (4). Offloading high-pressure regions of the foot is an essential strategy to prevent the onset of diabetic foot ulcers and to treat active ones (5,6). This offloading requirement is usually addressed by specific therapeutic bespoke or off-the-shelf shoes, which are key components of ulcer prevention and management (7). It has been shown that specific footwear design features, such as rockers and rigid midsole/outsole materials, are effective in reducing pressure and re-ulceration rates (8,9).

Evaluating the isolated effects of individual footwear features is crucial for clinical research and helps promote a shift from experience-based methods to a more data-driven approach in footwear manufacturing (10). However, footwear-related studies often involve multiple design modifications simultaneously, such as variations in fit due to changing interventions, upper construction, last shape, and insole characteristics, thus making it difficult to determine cause-and-effect relationships between specific footwear features and observed biomechanical outcomes (3). This challenge is compounded by variability in study protocols and in the properties of intervention and control shoes, which can lead to biased or incomplete results and severely limit the strength of footwear design recommendations (11,12).

An approach to evaluate the effects of isolated footwear features on biomechanical parameters is to use modular or modifiable designs that allow the modification of a single feature while keeping the rest of the shoe unchanged. Cavanagh et al. (1997) developed an experimental modular extra-depth shoe that used the same upper and footbed while allowing different rocker-bottom modules to be tested (13). They studied the isolated effect of anteroposterior forefoot rocker-axis position on the pressure distribution under the metatarsal heads. More recently, a modular shoe with interchangeable heel components that can be attached to the same upper using screws has been proposed to test the effect of heel rocker parameters on plantar pressure distribution (14). Another approach to test the isolated effects of rocker settings used an experimental shoe with rails and sliders for continuous forefoot rocker adjustment (15). In another study, custom 3D-printed rocker midsoles with varying rocker radius and longitudinal bending stiffness were evaluated. The forefoot bending motion of the midsole was restricted with cable ties, ensuring that the observed pressure changes were attributable to outsole parameters rather than to differences in the upper or fit (16). While a few such experimental shoes have been proposed, to the best of the authors’ knowledge, no thorough validation - in terms of repeatability and accuracy of the measurements with respect to standard shoes - has been conducted.

To overcome these limitations, the present study had two main aims: 1) to develop a modular footwear setup (MFS) to test the isolated effects of different midsole solutions on biomechanical parameters, such as foot and lower-limb kinematics and plantar pressure distribution, and 2) to validate the MFS by assessing its reliability and accuracy with respect to a standard control shoe with the same-design. The core of the MFS is the fixation system, which was required to allow easy attachment/detachment of the midsole components to/from the upper and to provide stable fixation of the midsole during dynamic tasks. Additionally, the fixation system was designed to have minimal impact on biomechanical parameters compared to a standard shoe with the same design.

## Materials and Methods

### Population

Ten healthy adults (5 M, 5 F; age = 33.2±9.2 yrs; height = 1.73±0.11 m; weight = 65.0±14.1 kg; BMI = 21.5±2.8 kg*m^-2^) were enrolled in the study. The inclusion criteria were age between 18 and 65 years, having a shoe size of 43 or 37 EU, and the ability to provide informed consent and comply with the tasks. Exclusion criteria comprised lower-limb musculoskeletal issues or surgeries, diabetes, peripheral neuropathy, and foot deformities.

The study received approval from the local ethics committee (AVEC #647/2025/Sper/IOR). All participants were informed about the data collection process and signed informed consent forms to participate.

### The Modular Footwear Setup

The MFS was designed and manufactured in different sizes (37-45 EU) to test the effect of midsole/outsole materials and shapes on biomechanical parameters. The MFS comprises three main components: a comfortable upper from an off-the-shelf orthopedic shoe featuring a flexible inner-insole (Botero, Podartis, Italy); a collection of interchangeable midsoles; and a micro-hook-and-loop fastener layer (Velcro, US) to allow easy fixation of the midsoles to the shoe upper (Figure 1A). A custom alignment device was designed for accurate and repeatable midsole attachment to the upper (Figure 1A). The device comprises an 8 cm wide aluminum track fitted with two 3D-printed cunei (Stratasys F370, US) made from Acrylonitrile butadiene styrene (ABS) filament. One of the two cunei can slide and be locked at any point along the aluminum track, ensuring firm positioning of the midsole for attachment. The upper, worn by the participant, slides in and is guided along the cunei until it attaches to the midsole.

**Figure 1.**
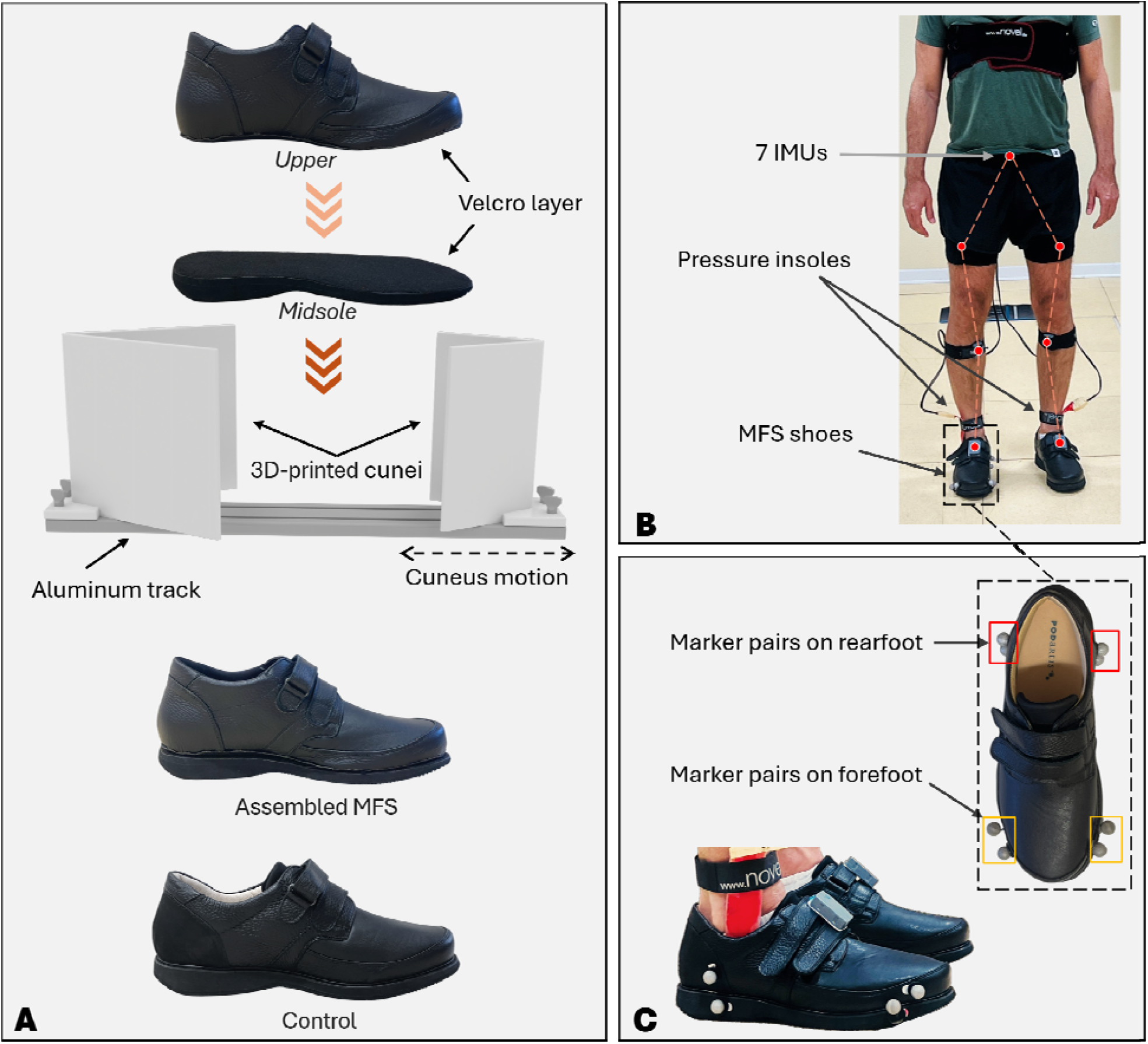
Brief graphical description of the experimental protocol. (A) Operation of the alignment device: the midsole is kept firmly in place by the two sliding cunei that can be adjusted and fixed along the aluminum track using screws. The participant wearing the upper gently slides the foot downwards guided by the cunei on top of the midsole. (B) An exemplary participant instrumented with seven IMUs and pressure insoles in the shoes. (C) Pairs of reflective markers attached to the shoes were used to measure the relative movement between upper and midsole during walking.

### Experimental Protocol

Participants were asked to walk at a comfortable walking speed along a 10 m walkway in two shod conditions: 1) wearing a standard comfortable orthopedic footwear (Botero, Podartis, Italy), and 2) wearing the MFS featuring an identical midsole. The order was randomized for each participant, and the walking speed was controlled using a stopwatch to be within 10% of the first walking trial. Five participants wore shoe size 43 EU, and five wore size 37 EU. They were asked to perform common tasks of daily living (e.g., walking, ascending/descending stairs, and tip-toe standing) to familiarize themselves with the shoes before data acquisition. Each participant was instrumented with seven Inertial Measurement Units (IMUs; Ultium Motion, Noraxon, US), sampling at 200 Hz. Four IMUs were attached to the shanks and thighs of the left and right legs using Velcro straps. One IMU was attached to the back just above the pelvis, and two were attached to the left and right shoes using double-sided adhesive tape (Figure 1B). IMUs were placed on the body segments and calibrated according to the manufacturer’s recommendations before each data acquisition. Temporal profiles of hip, knee, and ankle joint rotations in the sagittal and frontal planes were collected and normalized over gait cycle duration. Range of motion (ROM) was calculated for each joint as the absolute difference between the maximum and minimum angles in gait. Each shoe was fitted with a pressure insole featuring 99-capacitive sensors (Pedar X, Novel, Germany) sampling at 100 Hz ( Figure 1B). The following pressure parameters were recorded for each participant in each condition: mean and peak pressure [kPa] at the whole foot, rearfoot, midfoot, and forefoot regions of interest (ROIs). Data were analyzed using a custom software based on MATLAB (MathWorks, US). For each participant, biomechanical parameters were averaged over 5 gait cycles in each condition.

In addition, each participant completed a Visual Analogue Scale (VAS) questionnaire to assess the overall perceived comfort, walking stability, and comfort at push-off for each shoe condition.

### Statistical Analysis

Plantar pressure data were checked for normality using the Shapiro-Wilk test. Due to the small sample size (n=10) and the non-normal distribution detected in several parameters, paired Wilcoxon Signed-Rank Tests were used to assess possible differences in biomechanical parameters between the MFS and control shoes. A Bonferroni correction was applied to the significance level to account for multiple paired comparisons (α=0.0125) of plantar pressure assessment. Statistical Parametric Mapping (17) was used to assess differences in the temporal profiles of joint rotations between the two shoe conditions. The VAS data were analyzed using the Wilcoxon Signed-Rank test.

### Repeatability analysis

To assess the repeatability of biomechanical measurements when wearing the MFS, pressure and kinematic data were recorded for each participant across three sessions. At the end of each session, the IMUs were detached from the body segments, the pressure insoles were removed from the uppers, and the midsole components were detached from the uppers. Before each new session, the participant was re-instrumented with the IMUs, and the pressure insoles were inserted in the MFS uppers. The midsoles were re-attached to the upper via the alignment device.

Repeatability of pressure parameters across sessions was quantified using the Intraclass Correlation Coefficient (ICC 3,1), the Standard Error of Measurement (SEM), and the Coefficient of Variation (CV). Repeatability in joint ROM measurements was assessed via CV. All analyses were performed using MATLAB (R2025a, MathWorks, US).

### Analysis of the midsole-upper fixation strength

To assess the stability of the fixation between the MFS midsole and upper, four pairs of spherical reflective markers were attached to the midsole and upper at the rearfoot and forefoot (Figure 1C). Markers’ trajectories were tracked via an 8-camera motion analysis system (Vicon Motion Systems Ltd, Oxford, UK) sampling at 100 Hz. The temporal profile of the relative distance between two markers placed on the midsole and upper of each pair was calculated over three gait cycles in a sub-sample of five participants in both MFS and control shoe conditions. For each pair of markers, the Root Mean Square Error (RMSE) of the distance [mm] was calculated with respect to that in the static position.

## Results

This study evaluated the performance of the MFS across several domains: biomechanical comparability to a control shoe, inter-session reliability, mechanical fixation stability, and user experience. Figure 2 is reporting the assembled hip, knee, and ankle joint rotations in the sagittal and frontal plane over the normalized gait cycle in the MFS and control shoe conditions. SPM analysis did not show statistically significant differences in temporal profiles across any time frame.

**Figure 2.**
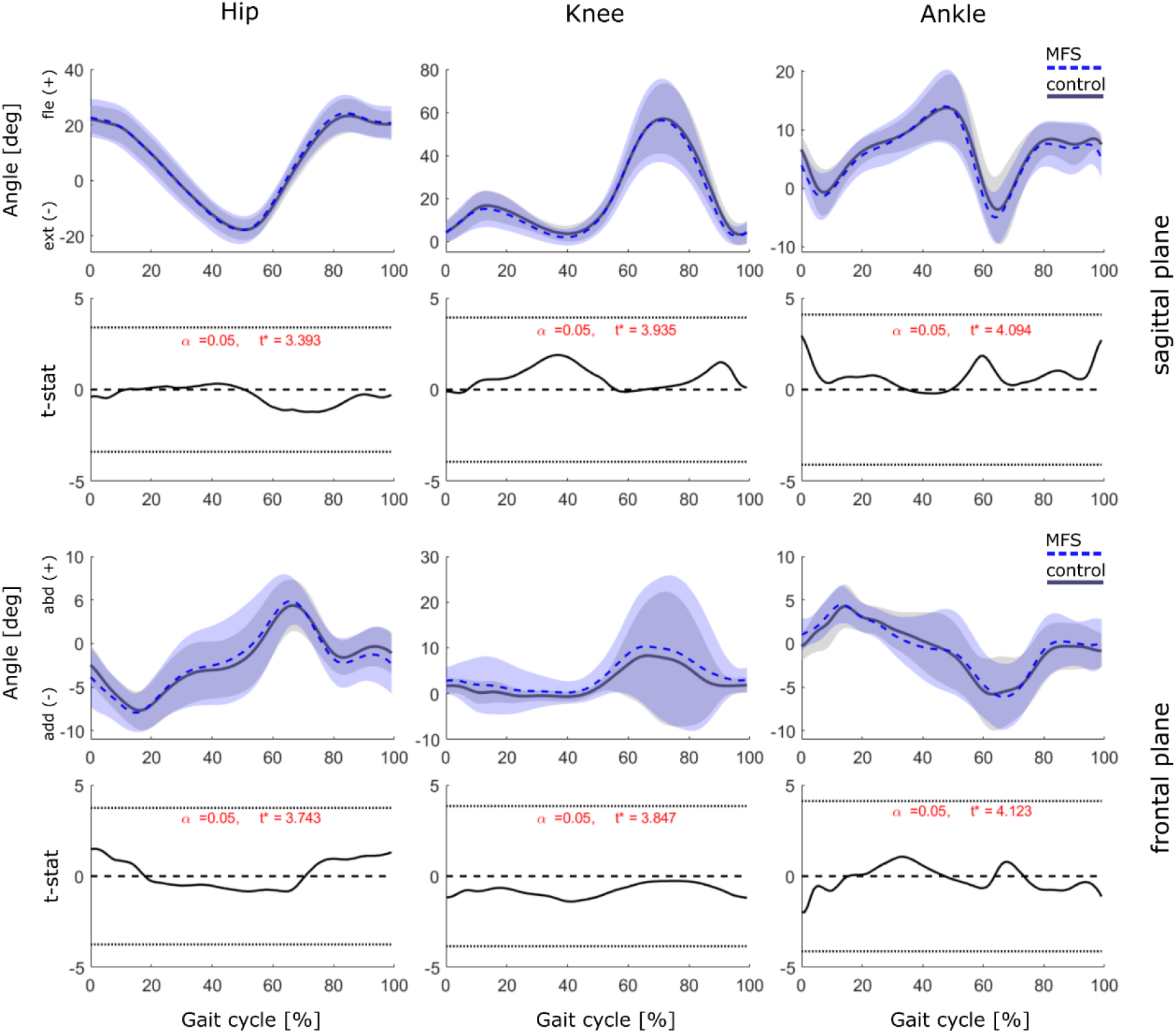
Temporal profiles (±1SD) of hip, knee and ankle joint rotations in the sagittal (top) and frontal (bottom) plane over normalized gait cycle in the MFS and in the control shoe. Statistically significant differences between shoe conditions have been investigated via Statistical Parametric Mapping analysis.

The inter-subject distribution of pressure parameters in all ROIs is summarized in Table 1. No statistically significant differences in pressure parameters were observed across ROIs between shoe conditions, except for rearfoot peak pressure, which was slightly lower in the control shoe.

**Table 1.**
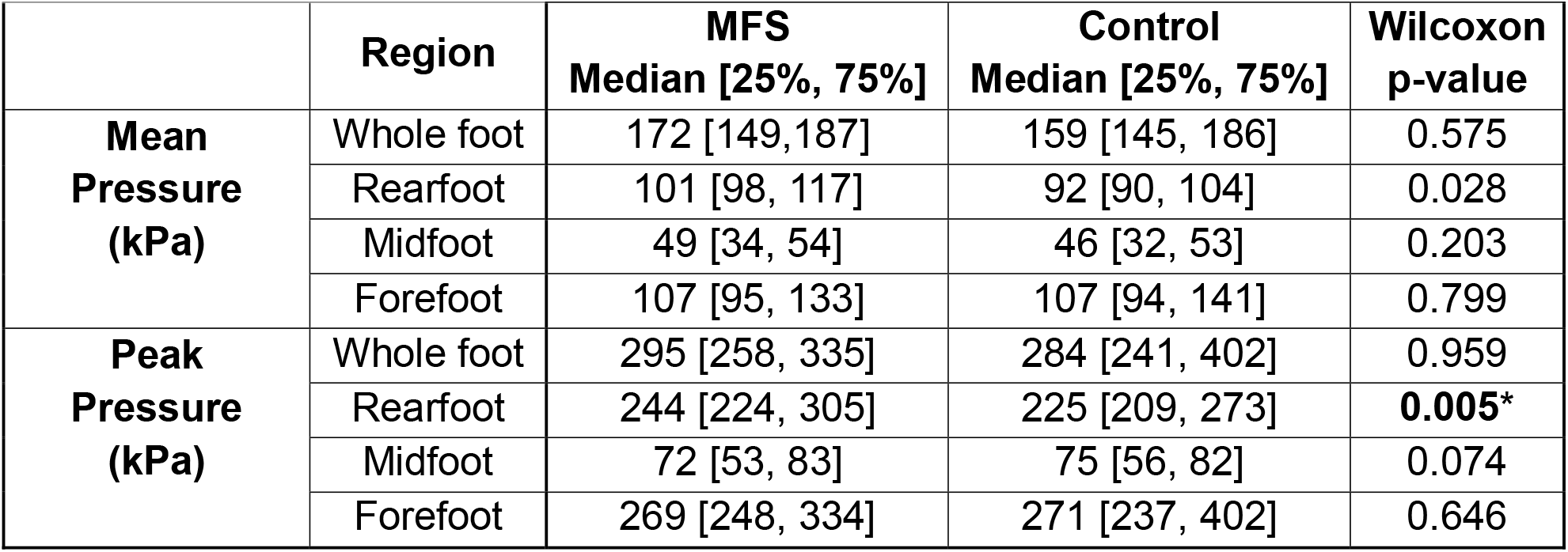
Inter-participant mean and peak pressure distributions in the MFS and in the control shoe in four plantar ROIs. Asterisks indicate statistically significant differences between shoe conditions (α=0.012).

|  | Region | MFS<br>Median [25%, 75%] | Control<br>Median [25%, 75%] | Wilcoxon<br>p-value |
| --- | --- | --- | --- | --- |
| **Mean**<br>**Pressure**<br>**(kPa)** | Whole foot | 172 [149,187] | 159 [145, 186] | 0.575 |
|  | Rearfoot | 101 [98, 117] | 92 [90, 104] | 0.028 |
|  | Midfoot | 49 [34, 54] | 46 [32, 53] | 0.203 |
|  | Forefoot | 107 [95, 133] | 107 [94, 141] | 0.799 |
| **Peak**<br>**Pressure**<br>**(kPa)** | Whole foot | 295 [258, 335] | 284 [241, 402] | 0.959 |
|  | Rearfoot | 244 [224, 305] | 225 [209, 273] | **0.005\*** |
|  | Midfoot | 72 [53, 83] | 75 [56, 82] | 0.074 |
|  | Forefoot | 269 [248, 334] | 271 [237, 402] | 0.646 |

Pressure measurements demonstrated high repeatability across the three sessions in most ROIs (Table 2). Specifically, mean and peak pressure at the midfoot and forefoot showed excellent repeatability (ICC > 0.9). Mean pressure at the rearfoot showed good repeatability (ICC > 0.8). Similarly, the range of motion for lower-limb joints showed excellent repeatability across sessions (Table 3), with low CV for hip flexion (0.06), knee flexion (0.05), and ankle dorsiflexion (0.08).

**Table 2.**
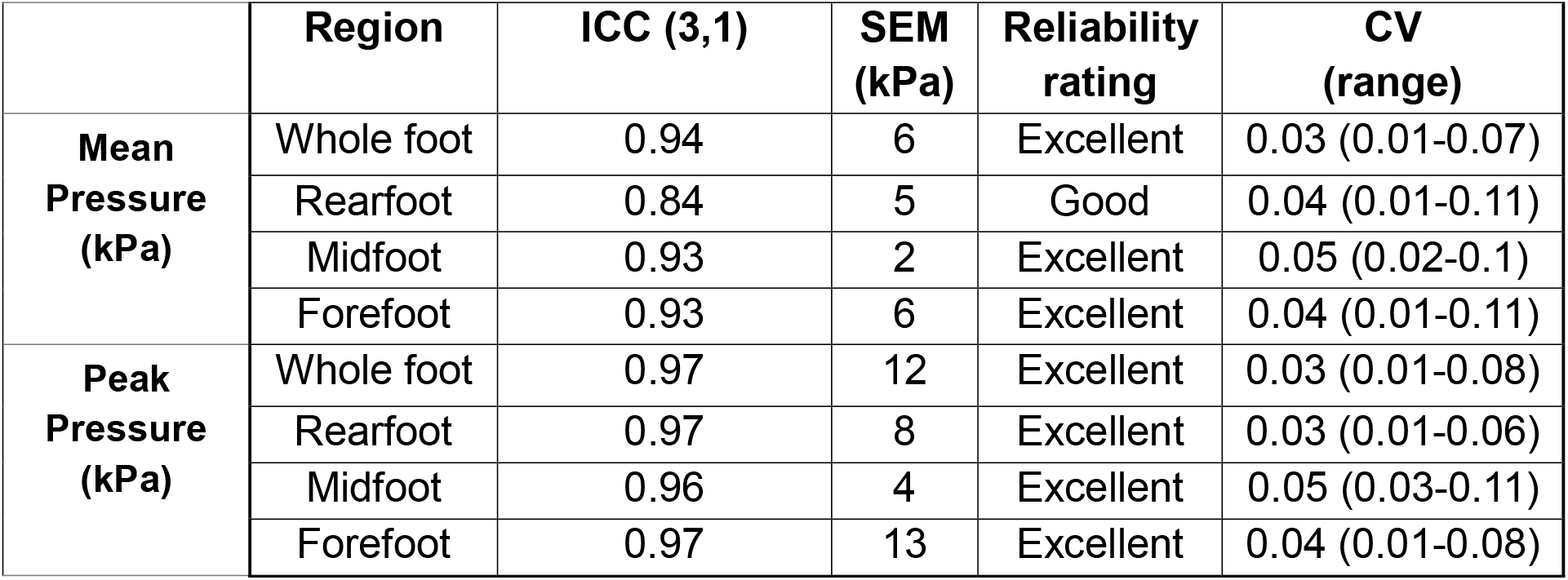
Repeatability of pressure measurements across three sessions wearing the MFS.

|  | Region | ICC (3,1) | SEM<br>(kPa) | Reliability<br>rating | CV<br>(range) |
| --- | --- | --- | --- | --- | --- |
| **Mean**<br>**Pressure**<br>**(kPa)** | Whole foot | 0.94 | 6 | Excellent | 0.03 (0.01-0.07) |
|  | Rearfoot | 0.84 | 5 | Good | 0.04 (0.01-0.11) |
|  | Midfoot | 0.93 | 2 | Excellent | 0.05 (0.02-0.1) |
|  | Forefoot | 0.93 | 6 | Excellent | 0.04 (0.01-0.11) |
| **Peak**<br>**Pressure**<br>**(kPa)** | Whole foot | 0.97 | 12 | Excellent | 0.03 (0.01-0.08) |
|  | Rearfoot | 0.97 | 8 | Excellent | 0.03 (0.01-0.06) |
|  | Midfoot | 0.96 | 4 | Excellent | 0.05 (0.03-0.11) |
|  | Forefoot | 0.97 | 13 | Excellent | 0.04 (0.01-0.08) |

**Table 3.**
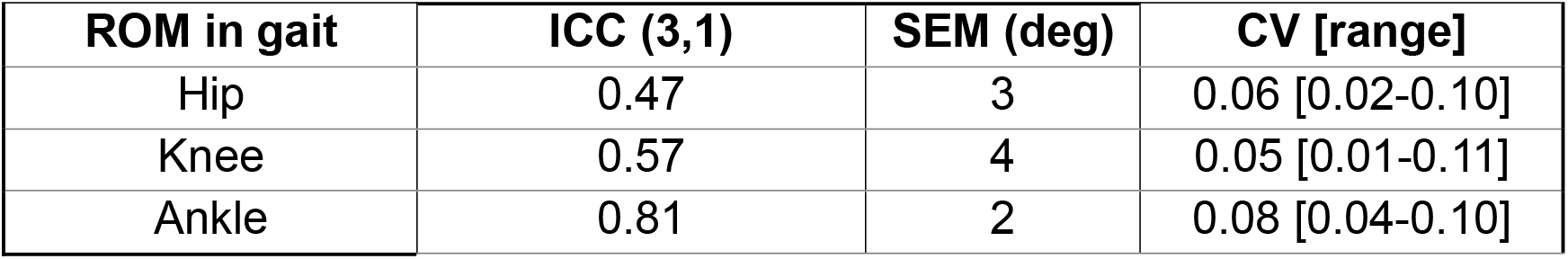
Repeatability of lower limb joint ROM in gait across three sessions wearing the MFS.

Analysis of the stability of the fixation between the MFS upper and midsole revealed very small relative displacements in gait (Appendix A). The RMSE of the displacement between markers in the rearfoot region of the MFS was slightly larger than that in the control shoe (1-1.5 mm vs. 0.5-1 mm), while no major differences were observed for the markers on the forefoot.

Participants rated the MFS slightly higher for overall comfort than the control shoe (Table 4). No significant differences were found regarding scores for stability or comfort at push-off between the two conditions.

**Table 4.** Outcome of the VAS-based perceived comfort, stability and push off in the MFS and in the control shoe. Asterisks indicate statistically significant differences between shoe conditions (α=0.05).

|  | **MFS score**<br>**Median (IQR)** | **Control score**<br>**Median (IQR)** | **Wilcoxon p-value** |
| --- | --- | --- | --- |
| Overall comfort | 77.4 (12.9) | 75.3 (13.7) | **0.023\*** |
| Stability | 75.3 (14.2) | 73.7 (9.7) | 0.065 |
| Comfort at Push-off | 78.9 (17.6) | 76.8 (12.6) | 0.797 |

## Discussion

The present study aimed to develop and validate a novel tool designed to isolate and analyze the effects of midsole modifications on lower-limb biomechanical parameters. The performance of the setup was evaluated across several domains: accuracy of biomechanical outcomes with respect to a control shoe, inter-session repeatability, mechanical fixation stability, and user experience.

The present MFS was developed through a trial-and-error process by testing several fixation solutions (see Appendix B). The aim was to create a tool that allows quick and repeatable midsole changes without affecting plantar pressure measurements relative to standard footwear. Earlier fixation designs (e.g., using 3D-printed custom pins or screws to attach the midsole to the upper) presented some limitations, including the time and effort required to change the midsoles, and the durability or strength of the fixation (see Appendix B). Alternative attachment methods (e.g. dovetail joints) were also considered; however, the discontinuity in the sole could affect plantar pressure measurements. Fasteners (18) and screws (14) to secure changeable midsole components, or cable ties (19) and cuts in midsoles (16) have also been reported to modify the mechanical properties of the midsoles without changing the upper. However, none of these methods were properly validated, thus making it difficult to determine whether the observed outcomes are solely due to the intended modifications or could be affected by the fixation method. Additionally, some of these designs are limited in the number of interventions that can be tested. For these reasons, a footwear setup using micro hook-and-loop fasteners was ultimately identified as the best solution.

According to the study’s findings, the MFS enables quick, strong, and reliable attachment of the midsole to the upper. Furthermore, the current setup allows pressure insoles and IMUs to stay in place while testing different midsole interventions, thereby enhancing measurement repeatability and reducing testing time.

The MFS provided a stable connection between the midsole and the upper during gait. Despite the flexible materials used for the uppers, the RMSE of the displacements between the upper and midsole components in the MFS during gait were in the range 0.5 - 1.5 mm and did not show major differences compared with the control shoe. None of the participants experienced midsole detachment during walking and in any of the dynamic tasks performed in laboratory conditions.

No differences in joint motion during gait were observed between the two shoe conditions (Figure 2). Additionally, the plantar pressure distributions and magnitudes while wearing the MFS closely resembled those seen in the control shoe, except for the peak pressure at the rearfoot. This small variation in this region may be explained by the fixation layer between the midsole and the upper, which could slightly increase the cushioning effect during the high-impact heel-strike phase. This was also evident from participants’ ratings, which indicated that the MFS was slightly more comfortable than the control shoe. However, there was no statistically significant difference in participants’ perceptions of walking stability and comfort at push-off. These results provide preliminary evidence that the current MFS design allows to replicate the biomechanical performance of a standard off-the-shelf shoe featuring the same midsole permanently attached to the upper.

The inter-session variability of plantar pressure parameters observed for the MFS is comparable to that reported for factory-made and conventional footwear (20–23). Across three sessions, both mean and peak pressure demonstrated good-to-excellent reliability (ICC (3,1) = 0.84–0.97). Thus, the MFS does not seem to introduce abnormal or excessive variability in pressure parameters across sessions. Indeed, the associated variability metrics remained comparable to those reported for standard footwear, suggesting that these differences reflect normal biological and gait-related fluctuations rather than instability of the shoe condition (20–22). Notably, the MFS appears to mitigate the regional variability often observed in studies reporting plantar pressure. While the midfoot is frequently identified as a region of lower repeatability in barefoot walking (ICC < 0.8 (22), and in clinical shoe measurements, where it often displays the highest variability (23), the MFS showed excellent repeatability in this region for mean and peak pressures (Table 2).

The current study provides strong evidence that midsole components evaluated using the MFS for plantar pressure, lower-limb joint kinematics, and comfort can accurately predict their performance in shoes produced via conventional assembling methods using adhesives or stitches. However, the results of the present validation study should be interpreted considering some limitations. First, only ten participants were recruited for this study, which may reduce the robustness of the results and increase the likelihood of false trends. Second, only walking trials in laboratory conditions were assessed for biomechanical parameters. While the micro-hook-and-loop fastener layer showed high attachment strength during gait, alternative midsole/upper fixation methods could be devised to provide a stronger connection for highly dynamic tasks such as running and other applications that require greater stability. Finally, only one upper condition was tested to evaluate the efficacy of the MFS. Testing and validating the connection strength across various upper designs and over longer durations could further broaden the applicability of this tool in footwear research.

## Conclusions

This study reports the development and preliminary validation of a new modular tool for testing the isolated effects of midsole designs on biomechanical parameters and perceived comfort. The tool could replicate the biomechanical performance of a standard off-the-shelf shoe featuring the same midsole and showed high repeatability for in-shoe pressure and kinematic measurements across sessions. The findings of the present study suggest the MFS has the potential to be a reliable and accurate tool for evaluating the effect of midsole features on relevant biomechanical parameters. This modular approach may enhance data-driven footwear design by providing a robust, simple-to-use, and replicable methodological framework to test the effect of midsole designs and materials for therapeutic, safety, and sport shoes.

## Data Availability

The data is available upon reasonable request.

## Acknowledgements

The authors would like to thank Filippo Goi for his help with producing the MFS parts, Anne Sturkenboom for her assistance in data collection, and Podartis SRL, Italy, for providing Botero shoes for this study. This project has received funding from the European Union’s Horizon 2020 research and innovation program under the Marie Skłodowska-Curie grant agreement no. 101073533 (DIALECT: Diabetes Lower Extremity Complications Research and Training Network in Foot Ulcer and Amputation Prevention). A part of this research was presented at DFSG 2025 as a conference poster.

## Disclosure

### Ethics approval and consent to participate

This study received approval from the local ethical committee (AVEC #647/2025/Sper/IOR). All participants were informed about the data collection process and provided informed consent to participate.

### Consent for publication

Not applicable

### Availability of data and materials

The data is available upon reasonable request.

### Authors’ contributions

K.S. and H.S. contributed to the conceptualization of the study, data curation, formal analysis, methodology, validation, and visualization, and were involved in writing the original draft and reviewing and editing the manuscript. G.R. helped with the methodology, provided supervision, validation, and review and editing. G.S. helped with the methodology, review, and editing. A.L. handled project administration, resources, supervision, validation, review, and editing. L.B. contributed to the project administration, supervision, review, and editing. P.C. was involved in conceptualization, formal analysis, methodology, supervision, validation, and the preparation, review, and editing of the original draft.

## Appendix A RMSE of the displacement of 4 marker pairs while walking, MFS vs Control

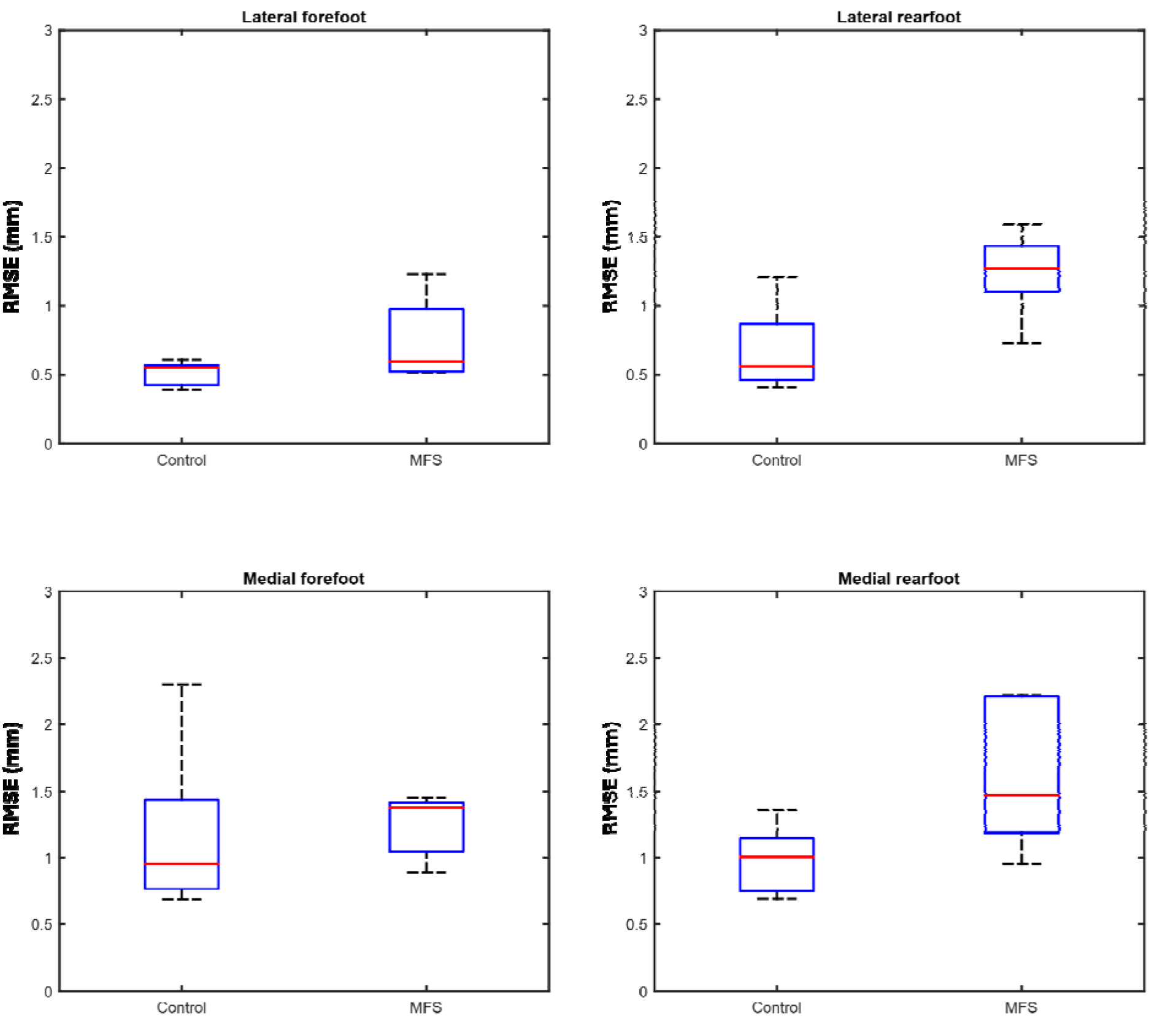

## Appendix B Selection of fixation system for the Modular Footwear Setup

The performance of each prototype is qualitatively rated relative to other design solutions. Rating in each category ranges from low to very high, where low is the lowest performance observed for the given prototype in that category, and very high is the best performance.

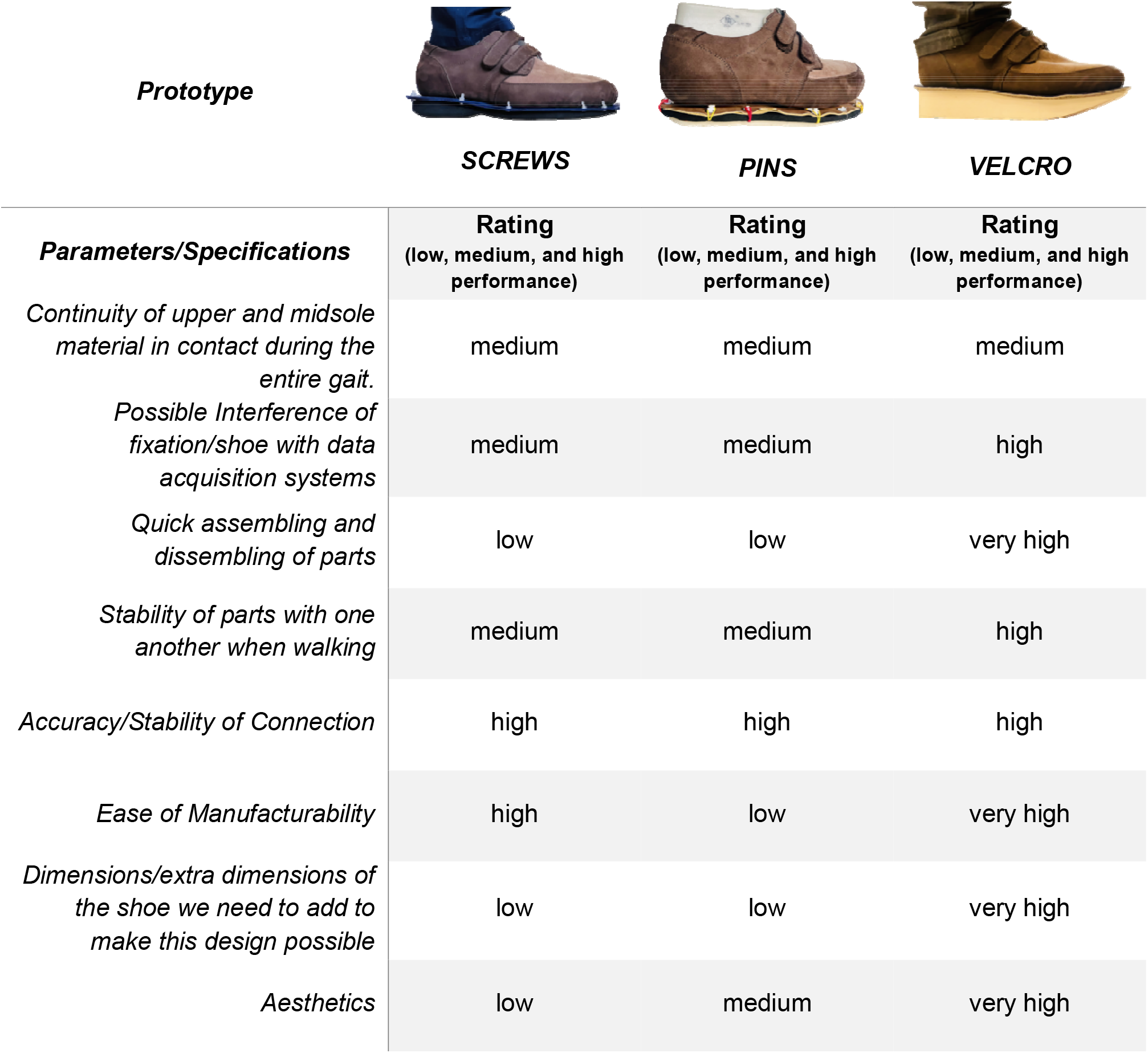

